# Prevalence, risk factors and clinical presentations of post-COVID-19 condition: a follow-up study of reported COVID-19 infections in Montreal, Canada

**DOI:** 10.1101/2023.12.15.23300022

**Authors:** Robert Allard, Safari Joseph Balegamire, Khadidja Boukar Malloum, Juliana Leal Ribeiro Cantalino, Céline Signor, Pascale Leclerc, Geneviève Bergeron, Sarah-Amélie Mercure, Geneviève Cadieux

## Abstract

**Background:** Meta-analyses suggest that post-COVID-19 condition (PCC) mostly takes three clinical presentations: fatigue, cognitive and respiratory. We sought to estimate the prevalence of these presentations and the strength of their associations with potential risk/protective factors, activity limitations and healthcare utilization.

**Methods:** Follow-up study by telephone re-interview, ≥5 months after initial interview, of a random sample of Montrealers aged 18 years or more, with a PCR-confirmed COVID-19 infection reported between July 18 and December 4, 2021. The re-interview covered age, sex, comorbidities, signs and symptoms (SS), activity limitations, healthcare utilization, perceived stigmatization and psychological distress.

**Results:** Of a sample of 2000 adults, 652 (39.1%) completed the questionnaire. Of 518 with only 1 acute episode of COVID-19, 32.2% met the WHO definition of PCC. Of these, 45.5% reported SS that fit the fatigue presentation, 24.6% the cognitive presentation and 16.8% the respiratory presentation. Neither age nor COVID-19 immunization was associated with PCC, compared to COVID-19 without PCC. However, being female (OR=2.28), hospitalization (OR=2.44) and intensive care (OR=3.21) for the acute COVID-19 episode were. Various activity limitations and types of healthcare utilization were also associated with PCC. All presentations were associated with serious psychological distress. The respiratory presentation was particularly associated with hospitalization for the acute episode (aOR=7.72) and was the only one associated with later hospitalization (aOR=23.6).

**Interpretation:** Our findings suggest that caring for patients with PCC requires adapted organizational models. If they favoured excellence in research, these models could help future studies meet the recommended methodological standards.

## INTRODUCTION

The prevalence of at least one COVID-19 sign or symptom (SS) persisting after an acute episode (with an average follow-up of 128 days), has been estimated at 37.8% (95% CI 31.8%-44.2%) in a meta-analysis of 36 studies.^1^ Thus, post-COVID-19 condition (PCC) appears to be a frequent occurrence after acute infection.

At the planning stage of this study, a meta-analysis^2^ of 41 studies reported the following as being significantly associated with PCC, in order of strength : hospitalization at time of acute episode, admission to an intensive care unit, being female, immunosuppression, chronic obstructive pulmonary disease, ischemic heart disease, age 40+, body mass index (BMI) of 30 or over, being a current smoker, and diabetes. Conversely, two-dose vaccination was strongly protective.

Another meta-analysis,^3^ of 54 studies, addressed the question of heterogeneity in the clinical manifestations of PCC and identified three clusters of self-reported SS: 1. persistent fatigue with bodily pain or mood swings, 2. cognitive problems: forgetfulness and difficulty concentrating (“brain fog”), 3. ongoing respiratory problems: shortness of breath with persistent cough. Their frequency was studied by age and sex, but not in relation to other potential risk factors for or effects of PCC.

Given the high proportion of the population that experienced acute COVID-19 and the relatively high reported prevalence of PCC among them, Montreal Public Health sought to better understand the determinants of the various clusters of SS of PCC and the effects of these clusters on Montrealers.

## METHODS

Eligible subjects were Montreal residents aged 18 or over with a PCR-confirmed SARS-CoV-2 infection reported to Montreal Public Health between July 18 and December 4, 2021, corresponding to the delta variant wave. As per routine case investigation procedures at the time, these individuals were interviewed by telephone soon after reporting. A random sample of 2000 were selected for reinterview, between May 12 and August 18, 2022, at least 5 months after their acute SARS-CoV-2 episode, by case investigators using a study-specific REDCap questionnaire in French or English, or in another language using simultaneous interpretation through *LanguageLine Solutions*.

Three attempts were made to reach the persons selected, at different times of day and on different days of the week, including weekends, between 8:30 and 18:30. Those not reached by telephone for whom we had a mobile phone number and/or an e-mail address were contacted through text messaging (SMS) and/or e-mail, asking for the best means and times to contact them. The questionnaire and interview procedures were piloted on 20 individuals. Exclusion criteria are listed in Figure 1.

**Figure 1.**
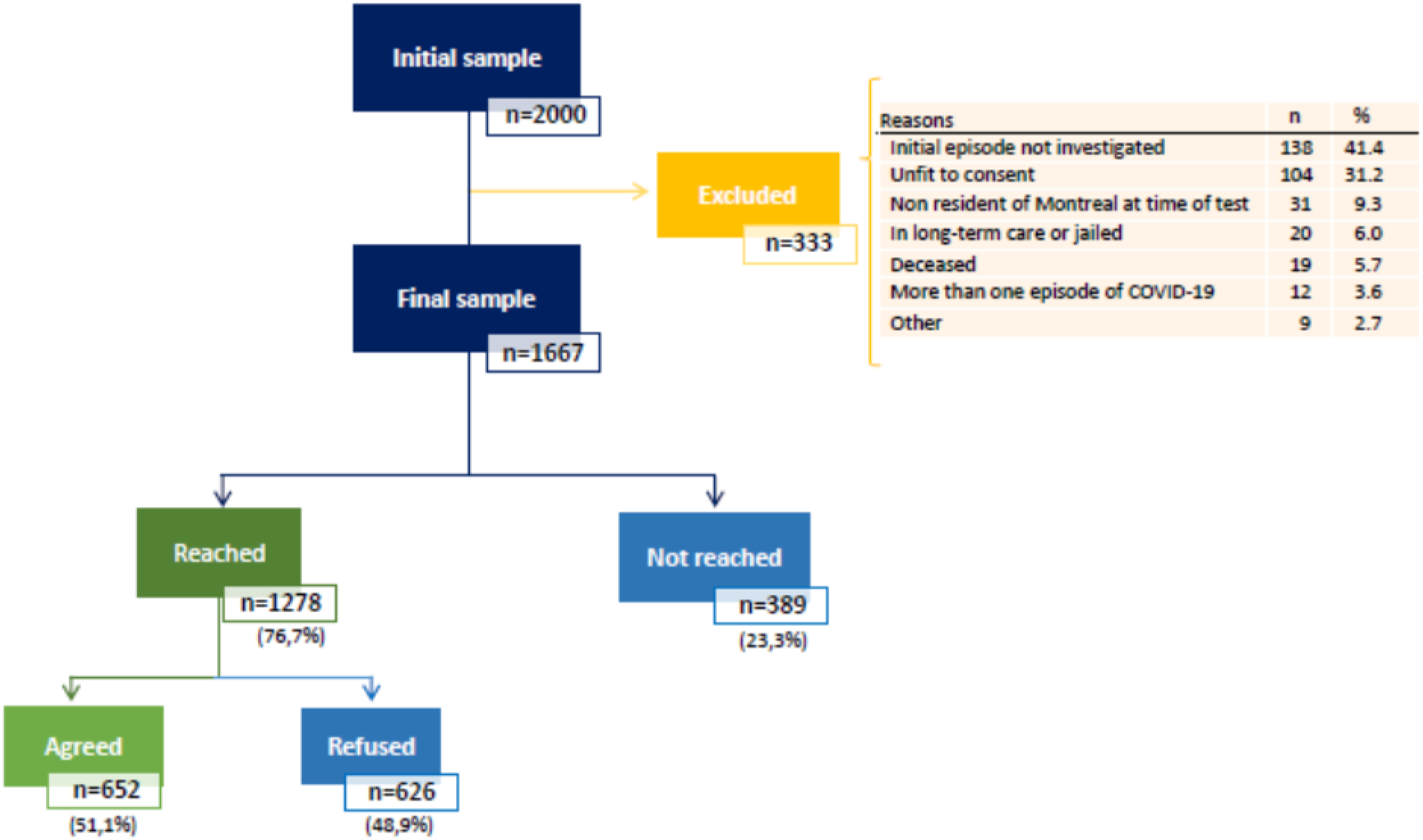
Flow diagram of the follow-up of the initial sample.

Basic socio-demographic characteristics were obtained during the initial COVID-19 case investigation and/or the study interview. Sex was self-reported as male, female or other. The study questionnaire included information about the acute episode, dates of onset and duration of 27 PCC-compatible SS and presence or absence of an alternate explanation for each, comorbidities, other potential risk/protective factors for PCC, and ethnocultural information. It also included questions on healthcare utilization (related to the SS reported) and activity limitations, both beginning at least 14 days after the acute episode, the Kessler-6 psychological distress scale^4^ for the month preceding the interview and the SSCI-8 stigmatization scale for a “recent” (as per the formulation of the question) period.^5^

Information on all COVID-19 vaccine doses received was obtained from the Quebec provincial immunization registry and matched with the questionnaire data using the unique provincial medical insurance number or, if missing, the last name, first name, sex and date of birth.

The definition of PCC used was that of the World Health Organization (WHO): “Post COVID-19 condition occurs in individuals with a history of probable or confirmed SARS-CoV-2 infection, usually 3 months from the onset of COVID-19 with symptoms that last for at least 2 months and cannot be explained by an alternative diagnosis.”^6^ The PCC *presentations* were adapted from the *clusters* in the recent meta-analysis,^3^ by replacing “and” in their definition with *and/or*, mostly because of the smaller sample size of our study. These were the *fatigue presentation*, defined by the presence of persistent fatigue and/or bodily pain (myalgia), the *cognitive presentation*, defined by the presence of forgetfulness (or difficulty concentrating, commonly called “brain fog”) and/or of memory problems, and the *respiratory presentation*, defined by the presence of shortness of breath and/or persistent cough.

Self-assessment of a change in general health status was measured by comparing the responses to the same question regarding two distinct time periods: before the acute episode and just before the interview. The difference score was categorized as improved, unchanged or deteriorated.

Twenty-four questions addressed activity limitations that began at least 14 days after the positive test. These related mostly to employment (14 questions), and the other questions to schooling/studies, personal care, activities of daily living, supporting others, social life, physical activity, leisure, money problems, and any other problem. Responses to individual questions were dichotomized as *all the time* or *most of the time* versus *sometimes, rarely* or *never*, when not already dichotomous. The overall score was the average, ranging from 0 to 4, of the actual scores for the questions answered by the respondent. This score was dichotomized as *≤1* versus >*1*.

The Kessler-6 Psychological Distress Scale was dichotomized based on a score of *13 or higher out of 24* versus *12 or less*, which represents “serious distress”^4^ during the 4 weeks preceding the interview. The 8-item Stigma Scale for chronic Illnesses (SSCI-8) was dichotomized based on a score *>8* versus ≤*8*, to indicate “recent”^7^ perceived stigmatization. Both these scales were grouped with activity limitations because they applied to the few weeks preceding the study interview, which makes them more likely to have followed than preceded the onset of PCC.

Each type of healthcare utilization was dichotomized as *at least once* since 14 days after the acute episode versus *never*.

Statistical analyses were performed using R software version 4.2.0, STATA version 15.1 and SPSS version 24.

In multivariate logistic regressions, variables that had a p value ≤ 0.15 in bivariate analyses were included in the initial models, which were refined using the likelihood ratio test and a backward stepwise selection method, with a significance threshold of 5% for retaining a variable in the final model. Four models, 1 for each of PCC and its 3 presentations, were tested for each grouping of independent variables (potential risk/protective factors, activity limitations, healthcare utilization). Age, sex and vaccination status were forced into all multivariate models, although their adjusted odds ratios (aOR) and 95% confidence intervals (95% CI) are presented in the tables only when statistically significant, as was done for the other aORs.

With respect to sample size, as a very approximate method, a sample size of 1200 was determined to yield sufficiently narrow confidence intervals in each of 4 age/sex subgroups with 300 respondents in each, for proportions with PCC ranging from 1% to 25%, based on estimates available at the time. An initial sample of 2000 would allow for 40% non-response.

The study protocol was reviewed and approved by the Research Ethics Board of the Montréal Regional Health Authority (project #2022-1582). Oral informed consent was obtained at the beginning of the study interview.

## RESULTS

Of the initial 2000 randomly selected subjects (Figure 1), 333 met the exclusion criteria. Out of the 1667 eligible ones, 1278 were reached and 652 agreed to participate in the study, for a response rate of 39.1% (652/1667).

Based on data obtained in the initial investigation, respondents were less likely than non-respondents to be younger than 30, compared to those aged 30 to 69 (OR=0.43 [95% CI: 0.31-0.60]) and less likely to be 70 or older (OR=0.58 [95% CI: 0.58-0.94]). Respondents were more likely to have been vaccinated against COVID-19 with 2 doses before their acute infection, rather than not vaccinated (OR=1.55 [95% CI: 1.22-1.96]). The two groups were otherwise similar on other variables available for comparison.

Among respondents, 134 were excluded because they had had another positive test result since their initial one, possibly representing a reinfection. This made it impossible to determine if they met the WHO definition of long COVID, there being more than one starting point to calculate the time intervals that are part of the definition. The following analyses are based on the remaining 518 respondents, who hereafter will be referred to as *participants*.

### Prevalence of PCC and its variants

Among participants, the 27 PCC-defining SS varied widely in reported frequency (Table 1) from 12.6% to 0.2%. PCC was present in 32.2% of participants (167/518; 95% CI: 28.4% - 36.4%), with 46.7% (78/167) showing only one SS, 15.6% two, 10.2% three, 8.4 % four and 19.1% between 5 and 15.

**Table 1.** Prevalence of reported PCC-defining SS, as formulated in the questionnaire. Denominators vary because of non-response.

| **Signs/symptoms** | **Numbers** | **%** | **Presentations** |
| --- | --- | --- | --- |
| Intense fatigue | 64/508 | 12.6 | Fatigue |
| Smell/taste altered or lost | 41/515 | 8.0 |  |
| Memory problems | 33/507 | 6.5 | Cognitive |
| Anxiety, nervousness | 28/511 | 5.5 |  |
| Muscle pain/cramps | 27/512 | 5.3 | Fatigue |
| Post-exertional malaise [explained in quest.] | 27/505 | 5.3 |  |
| Sleep problems | 26/514 | 5.1 |  |
| Shortness of breath or difficulty breathing | 20/514 | 3.9 | Respiratory |
| Joint pain | 20/508 | 3.9 |  |
| Brain fog, feeling like you cannot think clearly | 20/511 | 3.9 | Cognitive |
| Headaches | 18/511 | 3.5 |  |
| Dizziness, light-headedness | 18/511 | 3.5 |  |
| Vision problems, blurred vision | 17/513 | 3.3 |  |
| Depressive mood or depression | 15/511 | 2.9 |  |
| Heart palpitations, fast or irregular heartbeat | 14/509 | 2.8 |  |
| Menstrual period problems | 8/281 | 2.8 |  |
| Cough | 13/513 | 2.5 | Respiratory |
| Feelings of numbness or tingling | 13/513 | 2.5 |  |
| Loss of appetite or poor appetite | 12/501 | 2.4 |  |
| Tinnitus ringing or other hearing issues | 12/513 | 2.3 |  |
| Burning sensation or one as of an electric shock | 12/511 | 2.3 |  |
| New allergies | 9/510 | 1.8 |  |
| Chest pain | 9/515 | 1.7 |  |
| Other gastrointestinal problems [explained] | 6/505 | 1.2 |  |
| Stomach aches | 3/510 | 0.6 |  |
| Diarrhea | 2/510 | 0.4 |  |
| Fever | 1/506 | 0.2 |  |

Among the 167 participants with PCC, the fatigue presentation was reported by 45.5%, the cognitive presentation by 24.6% and the respiratory presentation by 16.8%. The 3 presentations together represented 60.0% of all PCC cases.

### Variables associated with PCC

Potential **risk/protective** factors significantly associated with a PCC on bivariate analysis (Table 2) were being female, reporting liver, immune system, inflammatory/joint or digestive problems, having been hospitalized or admitted to an ICU for COVID-19 within 14 days of the positive test.

**Table 2.**
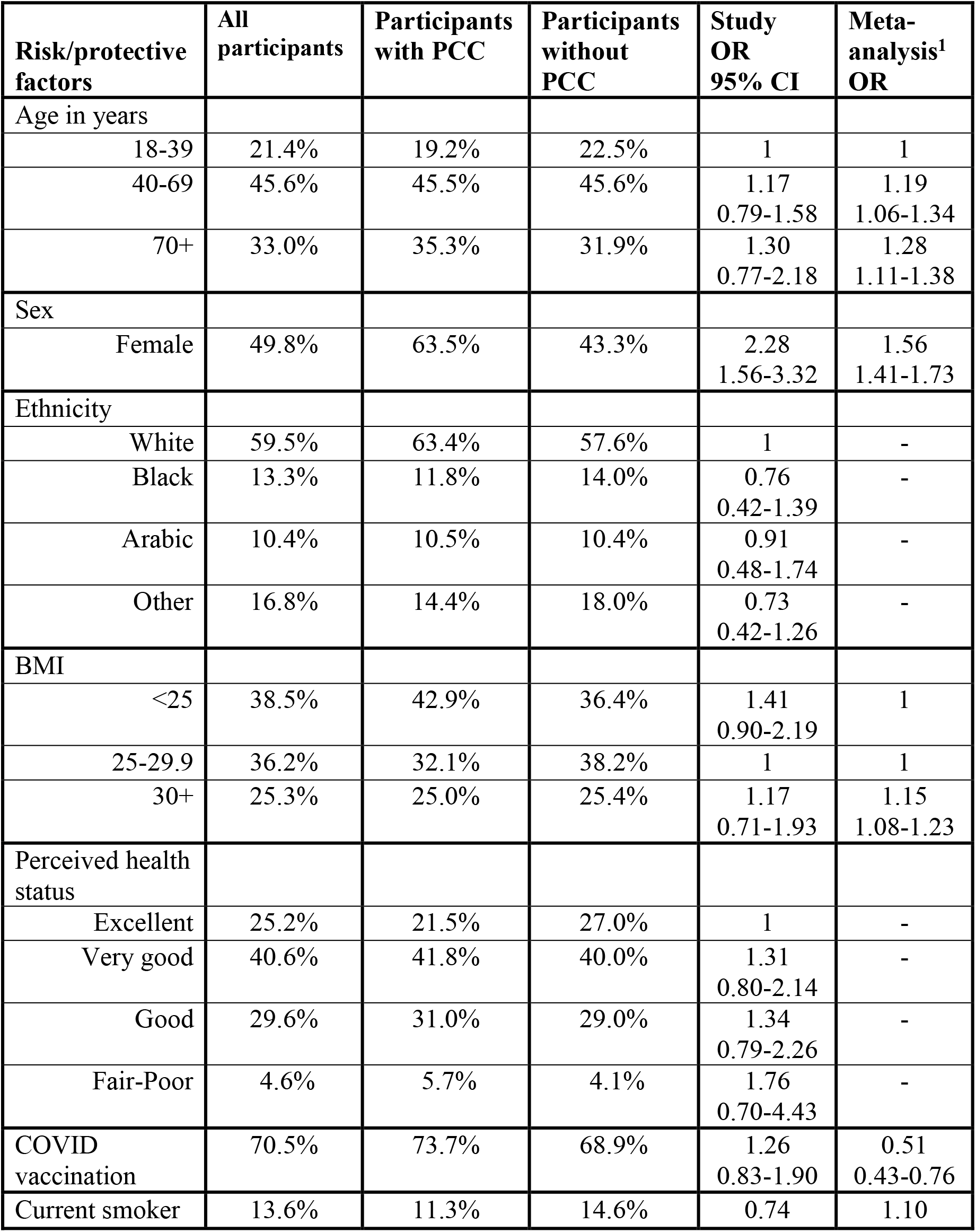
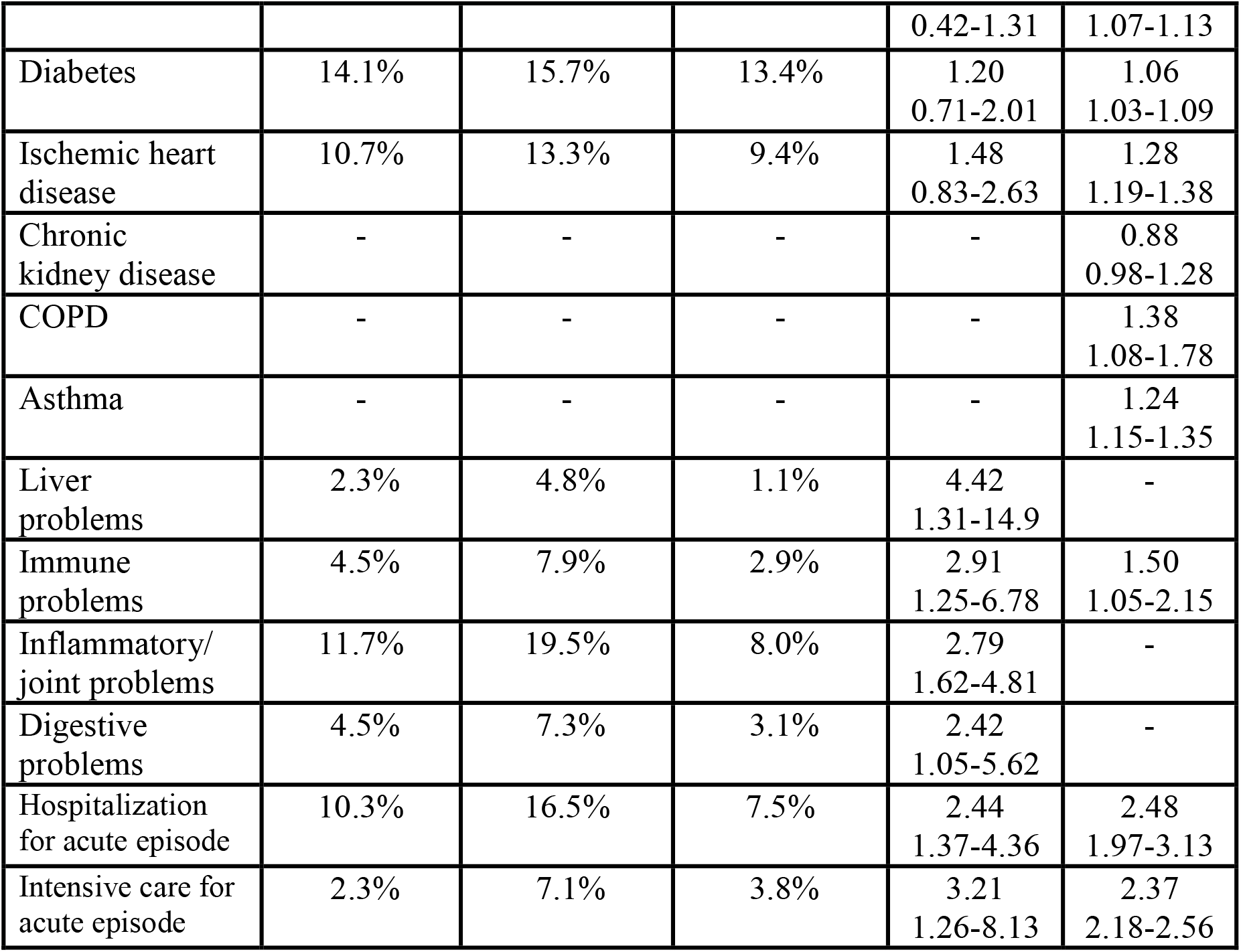
Prevalence of risk/protective factors among all participants and according to the presence or absence of PCC, with their unadjusted ORs, compared to the ORs from the meta-analysis of prevalence.^1^.

The reported **activity limitations** significantly associated with PCC on bivariate analysis are listed in Table 3. Notably, none of the work-related limitations were associated with PCC (data not presented), although all other types of limitations were strongly so.

**Table 3.** Prevalence and unadjusted ORs for activity limitations, overall health score after the acute episode and healthcare utilizations significantly associated PCC.

|  |  | All participants | Participants with PCC | Participants without PCC | Study OR<br>95% CI |
| --- | --- | --- | --- | --- | --- |
| Activity limitations | Physical | 13.3% | 28.8% | 6.4% | 5.80<br>3.25-10.3 |
|  | Leisure | 9.8% | 21.2% | 4.6% | 5.34<br>2.88-10.6 |
|  | Social | 11.0% | 21.5% | 6.3% | 4.08<br>2.26-7.35 |
|  | Routine | 6.8% | 13.5% | 3.8% | 3.97<br>1.93-8.15 |
|  | Personal care | 4.2% | 7.0% | 2.0% | 4.76<br>1.88-12.0 |
|  | Supporting others | 7.4% | 13.5% | 4.6% | 3.24<br>1.61-6.53 |
|  | Overall score | 17.4% | 32.5% | 10.6% | 4.04<br>2.81-6.52 |
|  | Perceived stigmatization | 29.3% | 37.1% | 25.6% | 1.71<br>1.15-2.54 |
|  | Psychological distress | 3.4% | 9.8% | 0.6% | 18.4<br>4.16-82 |
| Overall health score | Unchanged | 76.2% | 62.4% | 82.5% | 1 |
|  | Improved | 5.0% | 3.8% | 5.6% | 0.91<br>0.35-2.34 |
|  | Worsened | 18.8% | 33.8% | 12.0% | 3.72<br>2.33-5.94 |
| Healthcare utilization | Perceived need for consultation | 12.4% | 25.6% | 6.4% | 5.08<br>2.90-8.91 |
|  | GP consulted | 19.4% | 29.1% | 15.0% | 2.32<br>1.48-3.65 |
|  | Specialist consulted | 8.1% | 13.3% | 5.8% | 2.50<br>1.31-4.76 |
|  | Pharmacist consulted | 3.6% | 7.1% | 2.0% | 3.69<br>1.40-9.70 |
|  | Physiotherapist consulted | 1.6% | 3.8% | 0.6% | 6.81<br>1.36-34.1 |
|  | Massage Therapist cons. | 2.0% | 5.7% | 0.3% | 20.9<br>2.62-186 |
|  | Acupuncturist consulted | 1.6% | 4.4% | 0.3% | 16.0<br>1.96-131 |

A high perceived stigmatization score was reported by 29.3% of participants, that is, relatively frequently, but it had only a weak association with PCC (OR=1.71). Conversely, a high psychological distress score was reported by only 3.4 % of participants but it showed a very strong association with PCC (OR=18.4).

As for **healthcare utilization**, the most frequently consulted providers were family physicians (19.4% of respondents), specialist physicians (8.1%), nurses (4.0%) and pharmacists (3.5%). Respondents with PCC were significantly more likely to report consultations with family physicians, specialist physicians, pharmacists, physiotherapists, and especially massage therapists and acupuncturists.

Hospitalization after the acute episode was much more likely among those initially hospitalized: 25.0% (13/52) than among the others: 0.7% (3/452), OR=49.9 (95% CI: 13.6-183). The association was almost identical for those with and without PCC.

### Factors associated with the 3 presentations of PCC

Table 4 shows the potential risk/protective factors, health scores, activity limitations and healthcare utilization that were significantly associated with PCC and/or with any of its 3 presentations on multivariate logistic regression, adjusted for sex, age and vaccination status, and for each other within each of the above groupings of independent variables.

**Table 4.**
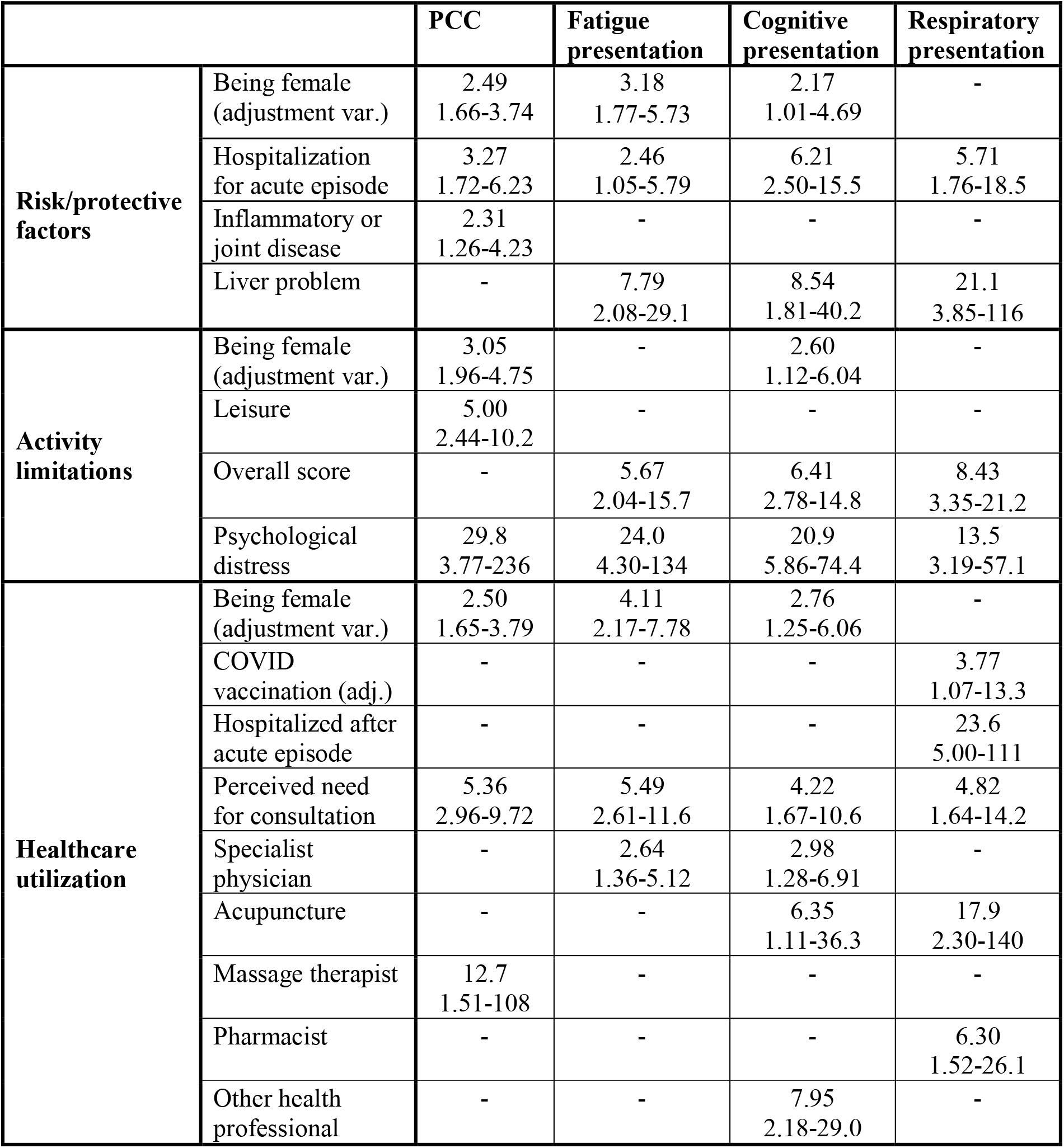
Associations of PCC as a whole and of its three presentations with risk/protective factors, activity limitations and healthcare utilization; odds ratios adjusted for sex, age and vaccination status.

Being female was associated with PCC globally and with its fatigue and cognitive presentations. Hospitalization for the initial COVID-19 episode, liver problems, the overall activity limitation score, severe psychological distress and perceived need for healthcare consultation were the variables most consistently associated with PCC. The fatigue and cognitive presentations were more strongly associated with psychological distress than the respiratory presentation, but the respiratory was the only one associated with hospitalization, during the acute episode and especially afterwards.

## DISCUSSION

In this study of 518 persons with PCR-confirmed COVID-19, 32% met the WHO definition of PCC, 60% of whom had either a fatigue, cognitive or respiratory presentation, or a combination of them. PCC, as compared to COVID-19 without PCC, was associated with being female, with several types of antecedent health problems and with hospitalization or ICU admission for acute COVID-19. The various PCC presentations were all associated with severe psychological distress and with a greater likelihood of consulting different types of healthcare providers. The respiratory presentation is particularly strongly associated with hospitalization after the acute COVID-19 episode; this can probably be explained by respiratory distress often requiring inpatient care.

A recent article discusses various limitations of the methods used to study PCC.^8^ It emphasizes the need for internationally established case definitions, clear inclusion and exclusion criteria, careful choice of control groups, proper matching of controls ? which can be interpreted more generally to mean better confounder control ? and sufficient follow-up time.

Our definition of *presentations* was more inclusive than that of *clusters* in the clustering meta-analysis^3^ and, as expected, our frequency estimates were higher. Several systematic reviews/meta-analyses have since then attempted to generate overarching clusters of SS. One, for instance, also arrived at three clusters “reached [by] a consensus through a discussion”: cardiorespiratory, inflammatory and neurological, corresponding very roughly to our respiratory, fatigue and cognitive manifestations.^9^ However, their inflammatory cluster is very heterogeneous, including, among other manifestations, hair loss, dizziness and sleep disorders. Another review, of 9 UK longitudinal studies,^10^ “identified symptoms characteristic of long COVID, including fatigue, shortness of breath, muscle pain or aches, difficulty concentrating and chest tightness”, again a set of SS rather similar to our presentations. The most recent review we could find^11^ identified fatigue, shortness of breath and cognitive dysfunction as the SS which present, 4 or more weeks after COVID-19 infection, the greatest risk ratio, compared in most studies to the risk in COVID-19-negative controls. These SS correspond exactly to the three manifestations we studied. However, there remains much disagreement about PCC profiles between studies and between systematic reviews.

Our response rate of 39% was lower than expected, but the respondents only differed from non-respondents as to age and vaccination status, among the variables available for comparison. These, and being female, were adjusted for in the multivariate analyses.

However, biases due to unmeasured confounders cannot be excluded.

The PCC prevalence estimate is vulnerable to participation bias, since subjects with PCC might be more likely to take part in follow-up investigations. However, it seems to us that this bias is unlikely to have affected differently those with different presentations of PCC, making comparisons between these more credible.

All recruitment occurred during the delta variant wave, possibly affecting the generalizability of the results to persons infected with other variants. Also, the frequency and nature of the PCC presentations might have been different had the patients been interviewed after another time interval than 5 months.

All comparisons were between persons who had acute COVID-19 infection. We have no information on the frequency of risk/preventive factors, activity limitations or healthcare utilization in the non-infected population. Many of the associations of PCC with potential risk factors that we observed had also been reported as significant in the meta-analysis of risk factors, the strongest ones being hospital/ICU admission, being female and immunosuppression^2^. The negative association with COVID-19 immunization^2^ was not replicated in our study. The most plausible explanation is that, in our study population, COVID-19 patients most at risk of PCC were also the ones most likely to have received the vaccine (confounding by indication), but that the resulting protection^12^ against PCC was insufficient to reverse this association.

## CONCLUSIONS

The high incidence of COVID-19 infection, the high prevalence of PCC among those infected, the strong association with serious psychological distress among them, the activity limitations they report and the variety of health services they seek, all bring out the need to care for patients with PCC using adapted organizational models. If these models favored research excellence and systematization, they could help future studies of PCC meet the recommended methodological standards.^8^

## Data Availability

The data used in this study cannot be made publicly available.

## ACKNOWLEDGEMENTS

The authors thank Jean-Philippe Proulx, BscInf, for his contribution to the supervision of data collection, all other Montreal Public Health staff who contributed to this study, including the COVID-19 case investigators, their supervisors and managers. Contributions: Conceptualization GB, GC, SAM. Data collection KBM, JLRC, CS. Analysis KBM, SJB, GC, RA. Interpretation and writing: RA, JLRC, SJB, KBM, PL, GB, GC.

The authors report no conflict of interest, financial or other, in relation to this work, which made use of already available public health resources and was not externally funded.

RA and SJB had full access to all the data in the study and take responsibility for the integrity of the data and the accuracy of the data analysis.

## Notes

### Competing Interest Statement

The authors have declared no competing interest.

### Funding Statement

This study did not receive any funding.

### Author Declarations

The Research Ethics Board of the Montreal Regional Health Authority gave ethical approval for this work (project #2022-1582).

